# Whole-Genome Long-Read Sequencing for Rapid Comprehensive Molecular Diagnostics of Brain Tumors

**DOI:** 10.64898/2026.04.23.26351563

**Authors:** Skarphéðinn Halldorsson, Richard Mark Nagymihaly, Christian Domilongo Bope, Marius Lund-Iversen, Pitt Niehusmann, Thomas Lien-Dahl, Jens Pahnke, Thomas Brüning, Geir Kongelf, Areeba Patel, Felix Sahm, Philipp Euskirchen, Henning Leske, Einar Osland Vik-Mo

**Affiliations:** Vilhelm Magnus Laboratory, Institute for Surgical Research, Oslo University Hospital, Oslo, Norway; Department of Pathology, Oslo University Hospital, Oslo, Norway; Division for Cancer Medicine, Oslo University Hospital, Oslo, Norway; INUM, University Medical Center Schleswig-Holstein, Lübeck, Germany; Department of Neuromedicine and Neuroscience, Faculty of Medicine and Life Sciences, University of Latvia, Riga, Latvia; Department of Neuroscience, Biochemistry and Biophysics, G.S.Wise Faculty of Life Sciences, Tel Aviv University, Israel; Department of Neuropathology, University Hospital Heidelberg, Heidelberg, Germany; German Cancer Consortium (DKTK), German Cancer Research Center (DKFZ), Heidelberg, Germany; Hopp Children’s Tumor Center (KiTZ), Heidelberg, Germany; Department of Neuropathology, Charité - Universitätsmedizin Berlin, Berlin, Germany; Department of Neurosurgery, Oslo University Hospital, Oslo, Norway; Institute for Clinical Medicine, Faculty of Medicine, University of Oslo, Oslo, Norway

**Keywords:** Molecular pathology, nanopore, whole-genome sequencing, CNS tumors, precision oncology

## Abstract

**Background:** Classification of central nervous system (CNS) tumors has become increasingly complex over the past decade, raising concerns about the availability, feasibility and sustainability of comprehensive molecular diagnostics. We have evaluated nanopore whole genome sequencing (nWGS) as a single workflow to replace multiple diagnostic assays.

**Methods:** We performed nWGS on DNA extracted from 90 adult CNS tumor samples (58 retrospective, 32 prospective) and compared the results to findings from standard of care (SoC) diagnostic work-up. Analysis was done through an automated workflow that consolidated diagnostically and therapeutically relevant genomic alterations, including copy-number variation, structural, and single-nucleotide variants, chromosomal aberrations, gene fusions and methylation-based classification.

**Results:** Nanopore WGS enabled final diagnostic classification in all samples with >15% tumor cell content, requiring ∼3 hours of hands-on library preparation, parallel sample processing, and sequencing times within 72 hours. Methylation-based classification was available within 1 hour and was concordant with the integrated final diagnosis in 89% of cases (80/90). All diagnostically relevant copy-number variations, single-nucleotide variants, and gene fusions were concordant with standard-of-care testing, and *MGMT* promoter methylation status matched in 94% of cases. In addition, nWGS identified prognostic and potentially actionable variants that were not reported or covered by SoC.

**Conclusions:** Nanopore WGS delivers comprehensive genetic and epigenetic results with a fast turn-around compared to standard methods. This enables efficient, accurate, and scalable molecular diagnostics of CNS tumors using a single platform. Its broad applicability supports its implementation in routine clinical practice and may be extended to other cancer types requiring complex genomic profiling.

## Introduction

The integration of molecular signatures in diagnosis has improved classification and prognostication of many tumors. This has come at the cost of increasing diagnostic complexity, often requiring extensive molecular profiling for accurate diagnosis, prognostication, and identification of therapeutic targets. Comprehensive genomic analyses, including the detection of small variants (SNVs and indels), copy-number variations (CNVs), and gene fusions are frequently required for final diagnosis. Additionally, DNA methylation profiling has become an essential tool in neuropathology [1, 2], and its use is also gaining recognition in other tumor types [3–5]. Several diagnostic platforms are often necessary to classify CNS tumors, including next-generation sequencing (NGS) of DNA and RNA, microarray analyses, pyrosequencing, and methylation arrays, partially supplemented by fluorescent in situ hybridization (FISH) [6]. Integrating these modalities into a cohesive pathological report is time-consuming and costly, frequently discouraging their full investigation [7]. This challenge is not unique to neuropathology, as extensive molecular characterization has demonstrated value in improving tumor classification, prognostication, and the identification of actionable variants across multiple cancer types [8–10]. Whole-genome sequencing (WGS) has been proposed as a potential all-in-one solution [8], but current short-read sequencing technologies do not detect methylation and struggle to resolve complex structural variants [11]. Nanopore sequencing is an established long-read sequencing technology that is a plausible alternative to short-read platforms due to its ability to detect epigenetic modifications from native DNA, while its long reads improve detection and characterization of structural variants [12]. Here, we evaluated nanopore-based whole genome sequencing (nWGS) for complete genomic profiling of CNS tumor samples. This approach was facilitated by an automated analytic pipeline and report generation that includes DNA methylation-based classification, *MGMT* methylation status, gene fusions, CNV, and small variants. We applied this workflow to 90 CNS tumor biopsies (58 retrospective and 32 prospective) and compared our findings to standard-of-care (SoC) molecular pathology to evaluate its diagnostic accuracy and clinical utility.

## Methods

### Patient Selection and Sample Acquisition

Patients ≥17 years of age undergoing surgery for suspected malignant CNS tumors at Oslo University Hospital with available fresh/ frozen material were enrolled following informed consent (REK 2016/1791). The study was approved by the regional ethics committee (REK 388359 and 853700). A retrospective cohort (n=58) and a prospective clinical cohort (n=32) were included (Table 1). Biopsy sites in the prospective cohort were selected based on preoperative multimodal MRI including 3D anatomical sequences like T1 gradient echo, FLAIR, T1 with gadolinium enhancement, as well as diffusion, SWI, perfusion, MR spectroscopy, and diffusion tensor imaging to identify the optimal site for biopsy. Intraoperative tissue sampling was performed under navigation (Brainlab) and, where appropriate, supported by intraoperative ultrasound to correct for brain shift.

**Table1:** Clinical characteristics.

|  | **Retrospective**<br>(n=58) | **Prospective**<br>(n=32) | **Overall**<br>(n=90) |
| --- | --- | --- | --- |
| **Gender** |  |  |  |
| Male | 32 (55%) | 19 (59%) | 51 (57%) |
| Female | 26 (45%) | 13 (41%) | 39 (43%) |
| **Age at surgery** |  |  |  |
| Range | 18-77 | 17-76 | 17-77 |
| Median | 51.5 | 48 | 51.5 |
| **Biopsy type** |  |  |  |
| Fresh | 0 | 31 | 31 (34%) |
| Frozen | 58 | 1 | 59 (66%) |
| **Tumor type** |  |  |  |
| Diffuse glioma, IDHwt | 38 (66%) | 12 (38%) | 50 (55%) |
| Diffuse glioma, IDHmut | 14 (24%) | 18 (56%) | 32 (36%) |
| Other | 6 (10%) | 2 (6%) | 8 (9%) |

### Standard-of-Care Diagnostic Workflow

Biopsy tissue was formalin fixed (PFA 4%) and paraffin embedded (FFPE). After histological evaluation, fluorescent in situ hybridization (FISH), multiplex ligation-dependent probe amplification (MLPA), pyrosequencing, Sanger sequencing and/ or NGS sequencing using Oncomine Childhood Cancer Research Assay (OCC panel, Thermo-Fisher) or TruSight™ Oncology 500 (TSO500 panel, Illumina) was performed to screen for clinically relevant molecular alterations that support the histology based differential diagnosis. FISH was performed on FFPE tumor sections using dual-color probes targeting *CDKN2A* (9p21) and CEP9 (Vysis/Abbott Molecular, IL, USA). Homozygous deletion was defined as complete absence of *CDKN2A* signals in ≥20% of nuclei with retained CEP9 signals. 1p/19q status was assessed by MLPA using the SALSA MLPA probemix P088-D1 (MRC Holland, Amsterdam, Netherlands), following the manufacturer’s protocol. Data were analyzed using GeneMarker (v3.0.0) software. Hotspot mutations in *isocitrate dehydrogenase 1* (*IDH1*) (codon 132), *isocitrate dehydrogenase 2* (*IDH2*) (codon 172) and the *telomerase reverse transcriptase* promoter (*TERT*p) region (chr5:1,295,228–1,295,250, GRCh37/hg19) were analyzed by PCR and Sanger sequencing (BigDye Terminator v3.1; Thermo Fisher Scientific). Sequences were aligned to reference sequences (*IDH1*: NM_005896.4, *IDH2*: NM_2068.4 and *TERT*: NG_009265.1) and analyzed using Mutation Surveyor software (SoftGenetics). In cases of diagnostically insufficient results or further confirmation of the diagnosis, an EPIC methylome analysis was eventually performed and classified on the web-based server of the Heidelberg brain classifier v12.8 (www.molecularneuropathology.org). The MGMT Pyro kit (Qiagen) was used to establish *O6-methylguanine DNA-methyltransferase* (*MGMT*-promotor region methylation status in 69 cases, according to manufacturer‘s protocol and an established institutional cut-off ≥ 10% (Håvik et al., 2012). Cases with methylation levels between 8 and 10% were deemed borderline due to ambiguous reports in terms of response to temozolomide therapy (Hosoya et al., 2022; Quillien et al., 2016). Eight prospective cases were analyzed with EpiDirect® MGMT Methylation qPCR Assay (PentaBase) as SoC, with a cut-off for unmethylated versus methylated classification at 3% but samples with 2.5-5% were also classified as grey zone. Histological features and molecular findings were used for a final multilayered diagnosis according to standard diagnostic protocols and classified according to the 2021 WHO Classification of Tumours of the Central Nervous System (Louis et al., 2021).

### Nanopore Whole-Genome Sequencing

Genomic DNA was extracted from fresh or frozen biopsies using the DNeasy Blood & Tissue kit (Qiagen). Sequencing libraries were prepared with the NEBNext companion module (New England Biolabs) and ligation sequencing kit (SQK-LSK114, ONT). Libraries were sequenced on PromethION flow cells (R10.4.1, FLO-PRO114M) on a P24 sequencing device running Minknow (version 24.02.19, ONT), one library per flow cell for 68-80 hours. Flow cells were washed (EXP-WSH004, ONT) and re-loaded up to two times. Live basecalling was performed via Dorado (version 7.3.11, ONT) with the SUP model (dna_r10.4.1_e8.2_400bps_sup@4.0.1) and 5mC_5hmC methylation calling in the CG context and alignment to reference genome (hg38) with minimap2 (version 2.24-r1122). Reads with Q-score below 10 were omitted.

### nWGS analysis

Automated analysis was performed on aligned BAM files upon completion of sequencing with the *DIANA* pipeline [13]. Briefly, this workflow performs sequencing quality estimations, methylation-based classification via CrossNN (Yuan et al., 2025) and Sturgeon (Vermeulen et al., 2023), tumor-cell content (TCC) estimation, CNV analysis, reports gene fusion events, determines *MGMT* methylation status and calls and annotates small variants in user selected genes. In this study, small variants were called and reported in all genes included in the OCC panel. The workflow also produces input files for online methylation classification with MNP-Flex (https://epignostix.com/). CrossNN and MNP-Flex classifications at the methylation class level or for glioblastoma, IDH-wildtype (“GBM, IDHwt”) at the class family level were considered valid. As Sturgeon does not aggregate classifications to the class family level, all “GBM, IDHwt” classifications of any class were regarded as GBM, IDHwt.

nWGS data was not integrated into final diagnostic or clinical decision-making as part of the study.

### Benchmarking

Classifier and variant detection benchmarking was performed with the “caret” package in R. SoC analysis was treated as *Truth*, methylation classifiers were benchmarked against final diagnosis, variants (CNVs and SNVs) were benchmarked against reported findings by the relevant SoC platforms.

## Results

### Data generation

To optimize sequencing time and evaluate the analysis workflow, we retrospectively sequenced 58 brain tumor samples from frozen, archival biopsies. This yielded a median sequencing depth of 37.4x (18.1-51.6x, Figure 1A) and a median read length N50 of 11.15 kbp (4.8 kbp-16.6 kbp, Figure 1B). The median TCC among the retrospectively analyzed samples was 62% (9%-100%, Figure 1C) as evaluated by copy-number variation. Estimated TCC was >15% in 55 of 58 (95%) of the retrospective samples. TCC in 20 sequenced samples, as estimated with ACE, was compared to histomorphologically evaluated TCC in frozen sections prior to sequencing (Figure 1D). A standard operating protocol was established for weekly sequencing runs of DNA extracted from fresh or snap-frozen tumor tissue, sequenced for 72 hours followed by automatic report generation. In a prospective cohort of 32 samples, the median whole-genome coverage was 45.7x (29.1-58.2x), which was significantly higher than that of archival samples (Wilcoxon signed-ranked test, p=0.00015, Figure 1A). The biopsies in the prospective cohort were all neuro-navigated, with a median estimated TCC of 77% (18%-100%, Figure 1C) which was significantly higher than that of the retrospective cohort (64%, Wilcoxon signed-ranked test, p=0.033, Figure 1C).

### Methylation classification

We performed DNA methylation-based classification using three recently released classifiers: CrossNN with a published high-confidence cut-off of 0.2 [14], Sturgeon with a published high-confidence cut-off of 0.95 [15], and MNP-Flex with a published high-confidence cut-off of 0.3 for all platforms [16]. The CrossNN and Sturgeon classifiers are trained on the v11.4 DNA methylation reference dataset and do not include recently defined CNS tumor entities, such as diffuse paediatric-type high-grade glioma, H3-wildtype and IDH-wildtype (pHGG). The MNP-Flex classifier implements the updated Heidelberg v12.8 reference dataset encompassing the WHO 2021 CNS tumor entities. CrossNN and Sturgeon are specifically designed to accommodate sparse data. To the best of our knowledge, neither of them has been evaluated on nWGS data. To verify their utility in the context of nWGS, we ran all classifiers on the retrospectively sequenced samples. Samples with TCC < 15% (n=3) were not classifiable and were excluded from classifier evaluation. CrossNN had an accuracy of 0.946 (95% CI: 0.843-0.970) and Sturgeon 0.873 (95% CI: 0.737-0.915). The accuracy of MNP-Flex was 0.964 (95% CI: 0.875-0.996) (Supplementary figure 1). Threshold evaluation indicated that the published threshold of CrossNN (0.2) is also applicable to nWGS. However, the published thresholds of 0.3 for MNP-Flex and 0.95 for Sturgeon are overly conservative for nWGS data. Our data indicate that for nWGS data, the MNP-Flex threshold can be lowered to 0.2 and the Sturgeon threshold can be lowered to 0.9 without increasing misclassification rate. (Supplementary figure 2). Importantly, 53 of 55 samples (96.3%) had classification scores above the threshold of 0.2 for both CrossNN and MNP-Flex. In contrast, 36 of 55 samples (65.5%) had Sturgeon classification above the published high confidence threshold score of 0.95. Lowering the confidence threshold of Sturgeon to 0.9 resulted in 42 of 55 samples (76.4) reaching the threshold value (Supplementary figure 2). No misleading classifications were observed beyond this threshold.

**Figure 1.**
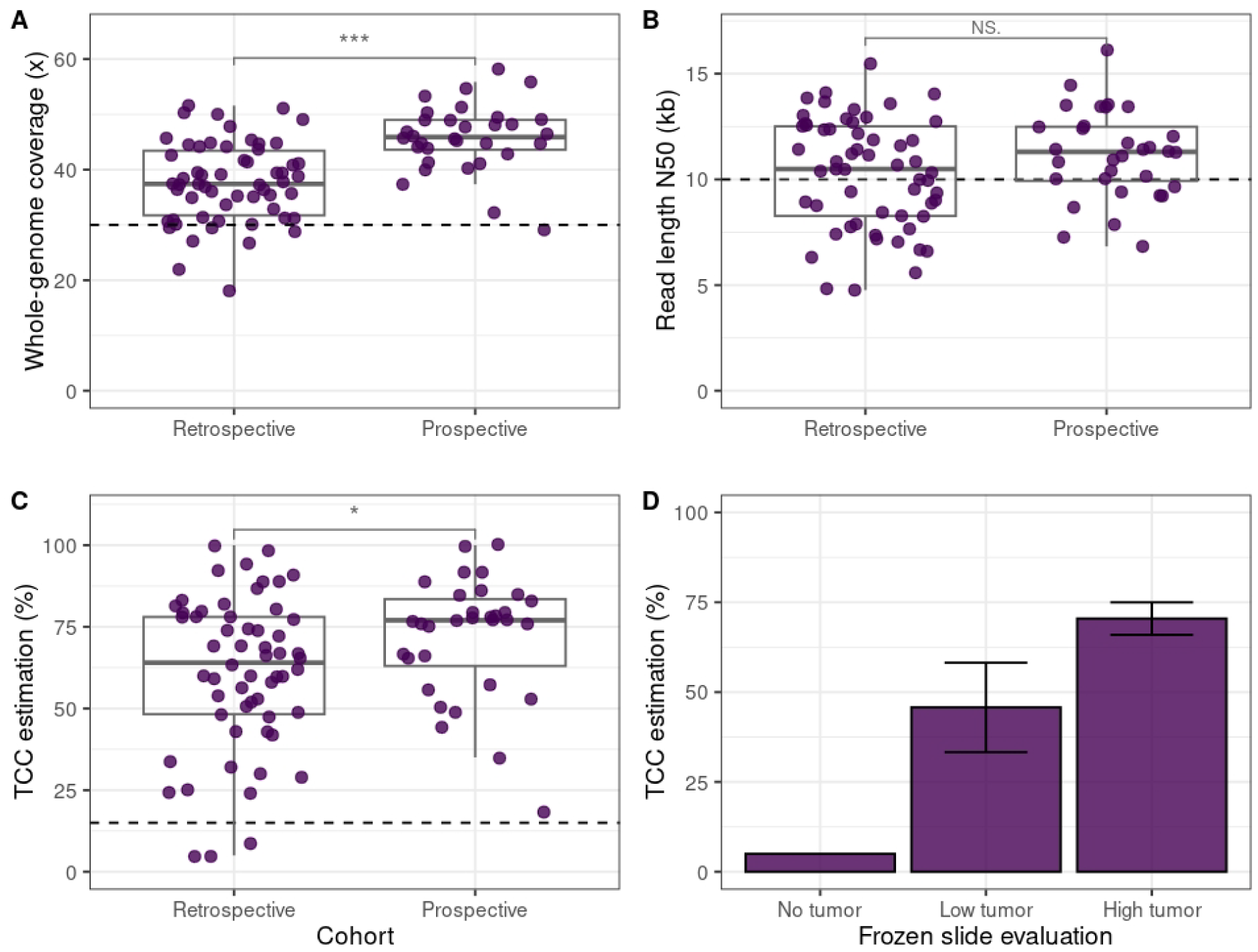
Sequencing metrics of all nWGS samples. **A)** Mean whole-genome coverage of every sample. The horizontal dotted line represents the predetermined target coverage of 30x. **B)** Read length (N50) of all samples. The horizontal dotted line represents the optimal read length (N50) of 10 kbp. **C)** *In silico* estimated TCC percentage of every sample. Horizontal dotted line represents the minimal TCC threshold (15%) compatible with analysis. **D)** Comparison of TCC as evaluated from frozen slides (n=20) prior to sequencing (x-axis) and *in silico* estimation following sequencing (y-axis). Error bars represent standard error. Cohort comparisons by Wilcox t-test, * = p<0.05, *** = p<0.001

All samples in the retrospective and prospective cohorts were assigned to methylation class and class family using the CrossNN, Sturgeon and MNP-Flex classifiers (Figure 3). CrossNN yielded classifications beyond control or inflammatory tissue (n=4) and with a score above the recommended threshold (score ≥ 0.2) for 81 of 90 samples (90%). The results were concordant with final diagnosis in 79 of 81 cases (98%). One ependymoma, posterior fossa type B (PFB), sample was classified as subependymoma, PF and a pHGG sample was classified as GBM, IDH-wildtype. Sturgeon yielded classifications above our determined threshold (confidence score > 0.9) for 71 of 90 samples (78.9%). Results were concordant with final diagnosis in 69 of 71 cases (97.2%). As with CrossNN, the ependymoma and pHGG samples were incorrectly classified.

MNP-Flex produced classifications above our determined threshold of 0.2 and beyond control or inflammatory tissue (n=3) in 80 of 90 samples (89%). Of these, 80 (100%) were concordant with the final integrated diagnosis. All classifier results and scores are provided in Supplementary Table 2. Three cases, classified by CrossNN and Sturgeon as “GBM, IDHwt”, “SUBEPN, PF” and “HGNET BCOR”, were refined by MNP-Flex as “diffuse pediatric-type high grade glioma” (pHGG), “Posterior Fossa Ependymoma (Subependymoma-enriched)” and “CNS tumor with EP300:BCOR fusion”, respectively. Six cases were evaluated by EPIC arrays as part of SoC (two GBMs, two high-grade astrocytomas with piloid features (HGAP), one pHGG and one CNS tumor with *EP300*::*BCOR* fusion). Results were concordant with the MNP-Flex classification in all these cases.

### Copy number variation

Relevant copy-number variations, including heterozygous gains/losses, homozygous deletions and focal amplifications were reported by the nWGS pipeline (Figure 2C, Figure 3). The pathognomonic 1p/19q-codeletion was detected in all 16 oligodendroglioma samples, in concordance with SoC evaluation by multiplex ligation-dependent probe amplification (MLPA). Gain of chromosome 7 and loss of chromosome 10 was observed in 32 of 48 GBMs (67%), and one astrocytoma (CNS WHO grade 3), IDH-mutant. Focal *EGFR* amplifications were detected in 23 of 48 GBMs (48%) and homozygous *CDKN2A/B* deletions in 31 samples (28 GBM samples (58%), two HGAP and one astrocytoma (CNS WHO grade 4), IDH-mutant. A *PDGFRA* amplification was identified in a pHGG, concordant with EPIC array based CNV analysis.

**Figure 2.**
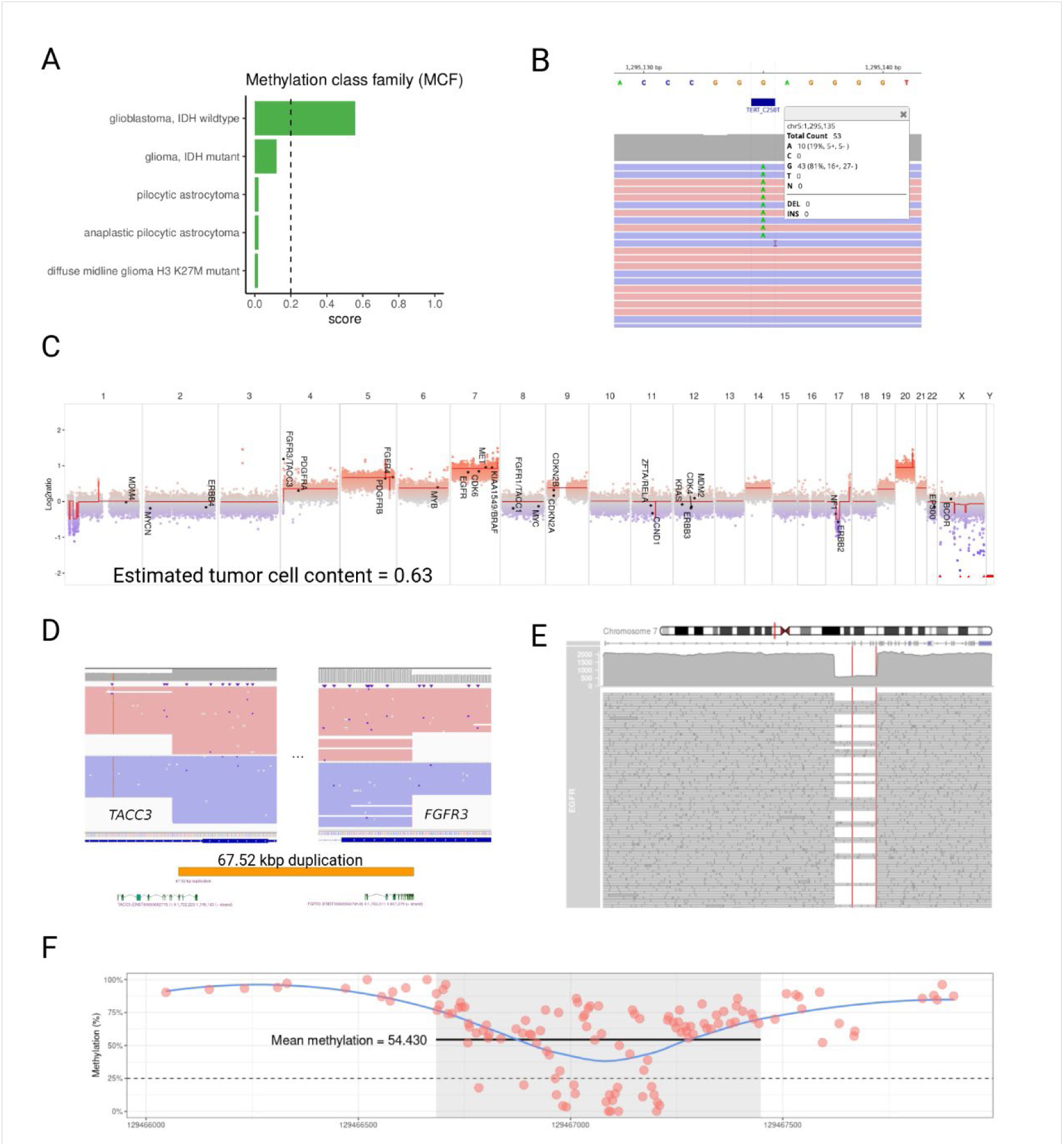
Examples of diagnostically relevant findings generated by nWGS. A) Results of methylation-based classification identify the sample as an IDH-wildtype GBM at the methylation class family (MCF) level. B) The sample harbors a C250T *TERT* promoter mutation (Depth=59, VAF=0.19). C) Copy-number variation plot shows multiple chromosomal aberrations including a gain of chromosome 7, loss of *ERBB2* (*Her2*) and a duplication involving *FGFR3* and *TACC3*. D) Structural variant analysis confirmed a duplication of a 67.5 kbp fragment on chromosome 4, causing a fusion of *TACC3* and *FGFR3*. E) IGV genome browser window of an *EGFR* amplification harboring a deletion that spans exons 2-7, causing an *EGFR*viii mutation. (Depth = 1899). F) Overview of the *MGMT* promoter methylation including all individual CpG sites in the *MGMT* CpG-island (grey box). Horizontal black line represents mean methylation on the CpG island, dotted line represents the threshold methylation value of 25% methylation that separates *MGMT* methylated and *MGMT* unmethylated samples.

**Figure 3.**
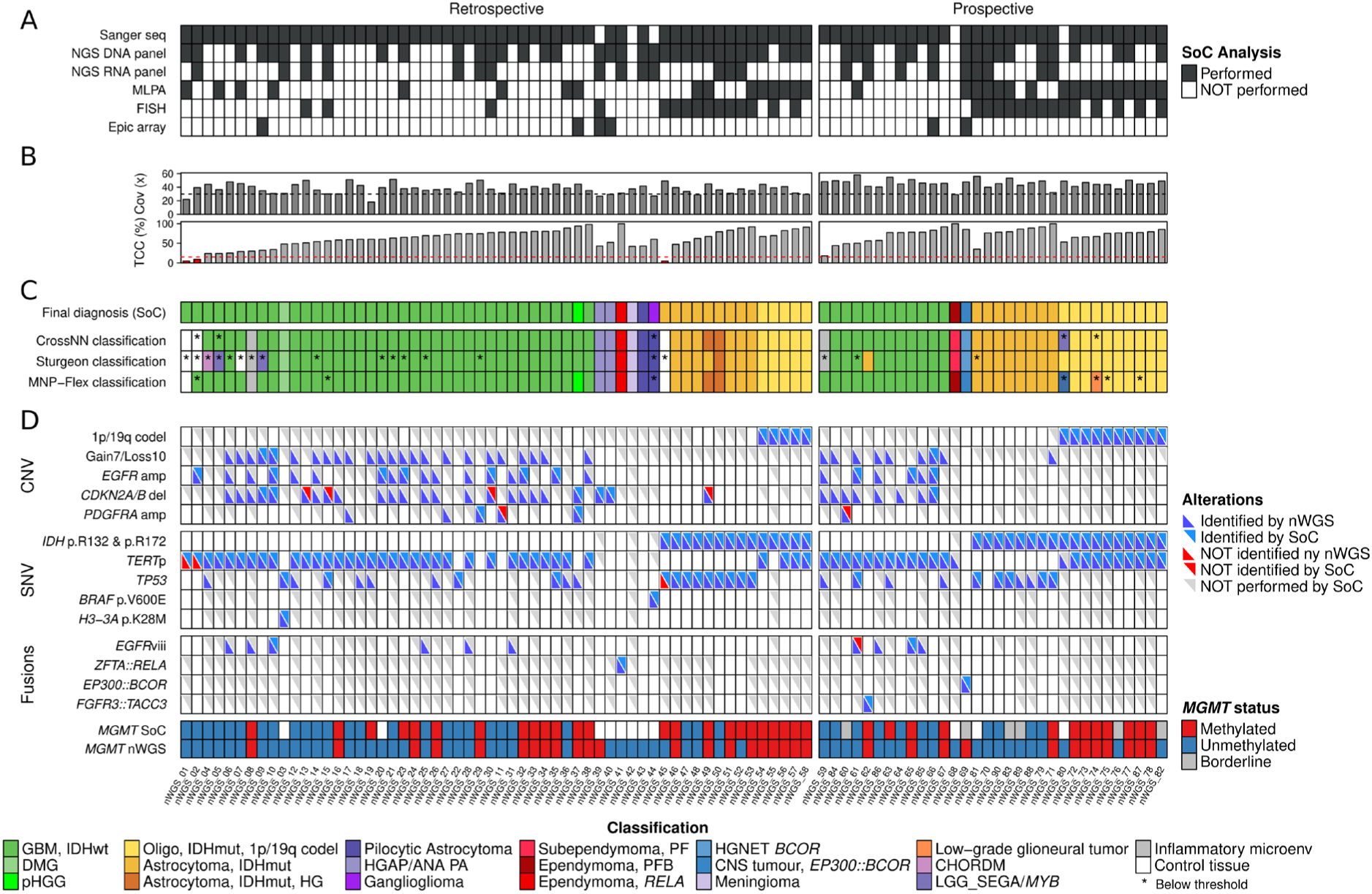
Summary of all analyses and diagnostically relevant findings in 90 CNS tumor samples. A) Analyses performed as SoC. B) Barplots demonstrating whole genome coverage (Cov) of samples after 72 hours of sequencing, dotted line marks the target coverage of 30x and estimated TCC percentage of samples based on copy-number profiling, red dotted line marks TCC of 15%, samples below not suitible for automated analysis. C) Classifier results for three methylation classifiers, CrossNN, Sturgeon and MNP-Flex. * represents classifier calls below threshold (< 0.2 for CrossNN and MNP-Flex, < 0.9 for Sturgeon). D) Overview of genomic alterations. Diagnostically relevant alterations of the cohort are listed and positive findings using nWGS and/or SoC are highlighted accordingly. MGMT methylation status as estimated by SoC (pyrosequencing or pentabase) and nWGS are shown.

Across 52 cases with available SoC data (NGS and/or EPIC Array), nWGS-based gene amplification status showed 100% sensitivity and 98% specificity. Additional amplifications (*EGFR, PDGFRA, CDK4, MDM2, MDM4* and *MYCN*) were reported via nWGS in 13 GBM samples lacking comparative SoC data. Homozygous *CDKN2A/B* deletion status, as reported by nWGS, was concordant with SoC (FISH/NGS/EPIC) in 21 of 25 samples where both methods were applied (sensitivity = 100%, specificity = 80%), with all mismatches reflecting nWGS based detection of homozygous deletions that were below the FISH probe resolution as described previously [17]. Nanopore-based WGS further reported homozygous *CDKN2A/B* deletions in 19 GBM samples where no *CDKN2A/B* related SoC data was available.

Gene amplifications with potential therapeutic implications as currently investigated in experimental studies [18] were reported in several GBMs by nWGS, including *PDGFRA* (n=4), *MDM4* (n=2), *CDK4* (n=3), and *MDM2* (n=2) where no SoC data was available for confirmation.

### Small genetic variants

As part of the nWGS workflow, small genetic variants (point mutations and indels) were called in all genes included in the OCC panel. Screening results for diagnostically relevant *IDH1* p.R132 and *IDH2* p.R172S alterations by nWGS were fully concordant with the SoC results (31 positive, 57 negative, 2 not investigated by SoC). Diagnostically relevant *TERT*p alterations were identified in 58 samples, absent in 30 and missed in two cases, both of which had very low TCC (Figure 3). Additionally, potentially pathogenic *TP53* variants were reported in 29 samples via nWGS (19 confirmed by SoC NGS, 10 not assessed by SoC). One *TP53* variant was missed in a sample with low TCC. A *BRAF* p.V600E variant was detected by nWGS in a ganglioglioma and an *H3-3A* p.K28M variant in a diffuse midline glioma, H3 K27-altered, both concordant with SoC results.

According to the recently updated EANO guidelines [18], various other genes may have prognostic or therapeutic relevance. SoC NGS reported 46 variants in 11 genes, 40 of which were also reported via nWGS (sensitivity 86.9%). All variants reported by nWGS in samples with NGS panel sequencing were confirmed (precision 100%). Overall, small variant detection in nWGS data showed 94% sensitivity (89% - 98%) and 100% precision (97% - 100%) compared to SoC (Table 2). Three of six false negative calls by nWGS were in samples with low TCC (<15%). All SNVs identified via nWGS or NGS in the cohort are listed in Supplementary Table 5.

**Table 2:** Varient detection benchmarking.

| **Variant type** | **Sensitivity (95% CI)** | **Precision (95% CI)** | **Specificity (95% CI)** | **Accuracy (95% CI)** |
| --- | --- | --- | --- | --- |
| CNV - all | **100%** (91.2 - 100%) | **83.3%** (70.4 - 91.3%) | **98.1%** (96.3 - 99.0%) | **98.1%** (96.3 - 99.1%) |
| CNV - exclude FISH | **100%** (91.2 - 100%) | **90.9%** (78.8 - 96.4%) | **99.1%** (97.6 - 99.6%) | **99.1%** (97.8 - 99.8%) |
| SNV- all | **94.2%** (89.3–96.9%) | **100.0%** (97.4–100.0%) | **100.0%** (99.5–100.0%) | **99.0%** (98.1–99.5%) |
| SNV- TCC > 15% | **96.0%** (91.6–98.2%) | **100.0%** (97.4–100.0%) | **100.0%** (99.4–100.0%) | **99.3%** (98.4–99.7%) |

### Gene fusions and structural variants

Diagnostically relevant gene fusions were detected by nWGS in three samples (Figure 2D, Figure 3, Supplementary Table 6). These comprised a *ZFTA*::*RELA* fusion in an ependymoma sample, an *EP300*::*BCOR* fusion detected in its new provisional entity and an *FGFR3*::*TACC3* fusion in a GBM sample. All three fusions were confirmed by NGS RNA panel sequencing. Four GBM, IDHwt samples showed additional alterations such as an *EGFR*::*SEPTIN14* fusion, a *NUP214*::*ABL1* fusion and two cases of *EGFR*::*EGFR* complex rearrangements without performed RNA panel sequencing for confirmation. Deletions of exons 2-7 in *EGFR* (*EGFR*viii variants) were detected in eight GBM samples, two of which were confirmed by SoC NGS panel sequencing.

### Methylation of *MGMT-*promotor region

*MGMT* methylation status as determined by SoC was available for 71 samples. *MGMT* methylation status was assessed based on 98 CpG sites in the CpG-island of the promoter region for all samples using nWGS data (Figure 2F, Supplementary Table 7). Dichotomous categorization of samples into methylated/unmethylated was concordant in 67 of the 71 samples (94%). In the four discordant samples, methylation-level according to SoC was 12-31%, which classified them as methylated (Figure 3). However, nWGS analysis of 98 CpG sites indicated low overall methylation (range 10-18%), the samples were therefore classified as unmethylated based on previous evaluation of full *MGMT* promotor methylation cut-offs [19, 20].

### Combined histological and molecular analysis

The nWGS workflow produced the diagnostically necessary data for complete molecular workup in all cases with sufficient TCC (Supplementary Table 1). In seven cases, methylation classifiers gave discordant results or no results above threshold. In five of these cases, histological evaluation as diffuse glioma in combination with the molecular findings produced via nWGS allowed for correct tumor classification. These comprised three GBM samples with IDH-wildtype status and at least one of the following alterations: gain of chromosome 7/loss of chromosome 10, *EGFR* amplification, and/or presence of a *TERT*p mutation. The other two cases comprised oligodendroglioma, IDH-mutant and 1p/19q-codeleted samples showing an *IDH1* p.R132H mutation, 1p/19q-codeletion and a *TERT*p mutation. In the case of a ganglioglioma (with detected *BRAF* p.V600E variant), none of the three methylation classifiers provided a classification above threshold so that histomorphological evaluation was decisive for the final classification. An ependymoma of the posterior fossa type B (PFB) was classified as “posterior fossa subependymoma” by both crossNN and Sturgeon but MNP-Flex produced a classification of “Posterior Fossa Ependymoma, Subependymoma-enriched”. This sample had no copy-number alterations but harbored a *TERT*p mutation, aligning it with subependymoma classification. However, the histomorphology was not consistent with a subependymoma, supporting the PFB ependymoma diagnosis; an inconsistency acknowledged in cIMPACT-NOW Update 11 (Wesseling et al., 2026).

### Sequencing time versus classification and variant detection

Sufficient sequencing depth is critical for robust and representative analytical results. The analytic tests necessary for CNS tumor diagnosis and prognostication require different sequence coverages. For example, the methylation classifiers CrossNN and Sturgeon were designed to accommodate very sparse data, where classifications are not expected to improve from 1x whole-genome coverage to 30x or beyond. In contrast, successful detection of somatic SNVs is a function of sequencing depth and TCC and will benefit from as much coverage as possible (Figure 4A).

**Figure 4.**
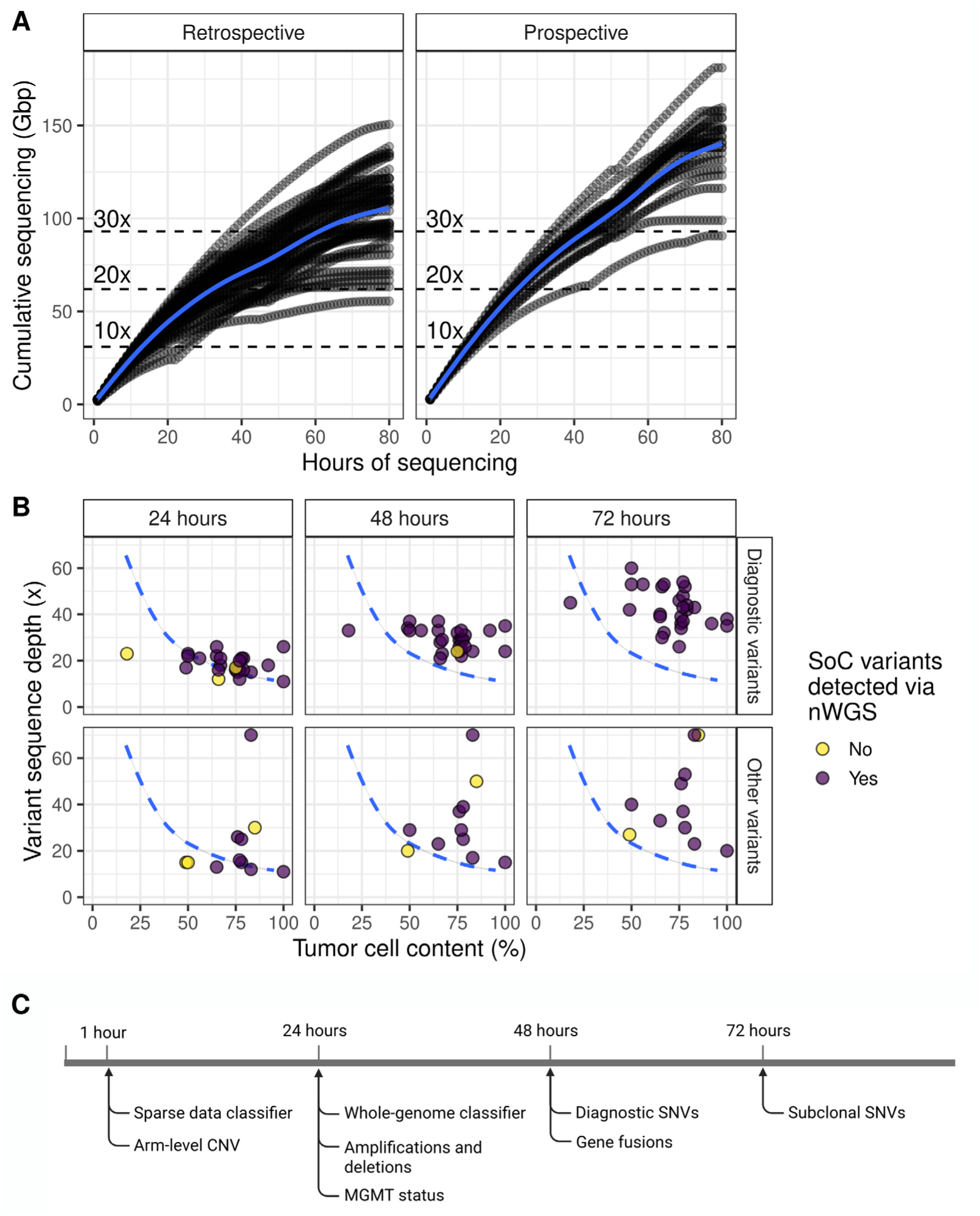
Sequencing output and variant detection limits. A) Cumulative sequencing output of all samples in the retrospective and prospective cohorts. Blue line represents the smoothed (LOESS) cumulative trend in each cohort. B) Detection of diagnostic and other relevant variants reported by SoC in the prospective cohort after 24, 48 and 72 hours of sequencing. The dotted blue line represents the theoretic 95% confidence threshold of capturing somatic variants at a given sequence depth and TCC. C) Timepoints when sufficient sequencing data for different analyses can be expected.

To evaluate the effects of various sequencing depths and sequencing time, we repeated all analysis on 20 samples in the prospective cohort with data generated after 1 hour, 24 hours and 48 hours of sequencing and compared the results to the final analysis after 72 hours of sequencing. Methylation classifications with CrossNN and Sturgeon were unchanged between 1 hour and 72 hours of sequencing for 18 of the 20 samples (90%). MNP-Flex cannot provide classifications after 1 hour of sequencing but provided 18 correct classifications after 24 and 48 hours of sequencing (90%) and 19 correct classifications after 72 hours (95%). Gross chromosomal alterations such as gain of 7/loss of 10 and 1p/19q codeletions, as well as focal gene amplifications, were correctly detected in all cases after one hour of sequencing. Homozygous *CDKN2A/B* deletions were reliably observed after 24 hours. *MGMT* methylation status was not reliably detected after one hour of sequencing, but identical results were achieved after 24, 48 and 72 hours of sequencing. Interestingly, 32 of the 38 (84%) diagnostic or pathogenic SNVs reported by SoC were accurately detected after 24 hours of sequencing and 36 of 38 (95%) after 48 hours of sequencing (Figure 4B). Live basecalling during nanopore sequencing allows analysis and interpretation at varying timepoints and sequencing depths if necessary (Figure 4C).

### Time and cost evaluation

The median turnaround time for the prospectively analyzed samples, from day of surgery to final report, was six workdays (two – eight workdays). The median turnaround time from day of surgery to final diagnosis for SoC was eleven workdays (five – fifty-seven workdays, Figure 5A).

**Figure 5.**
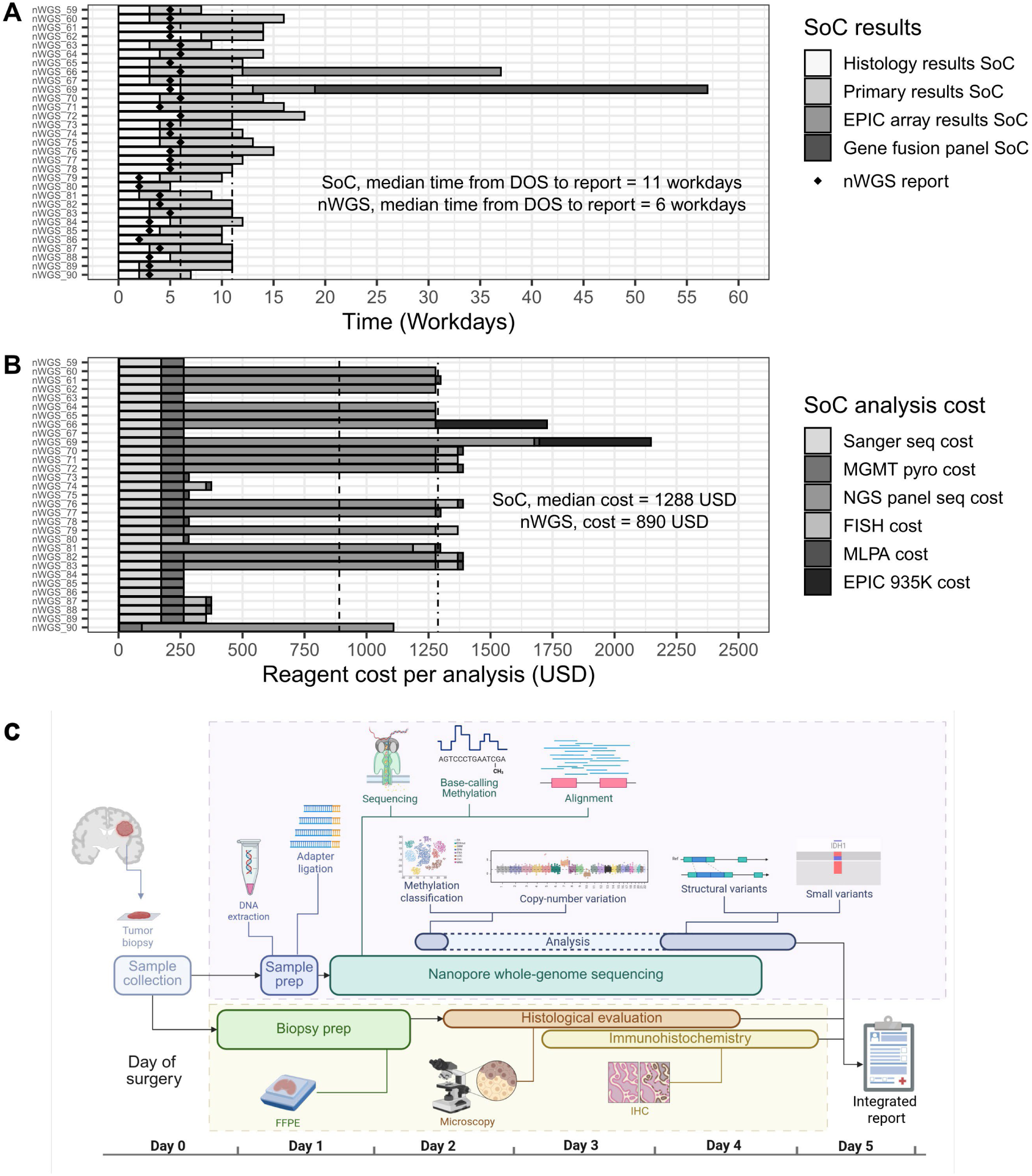
Turnaround time and reagent costs for nWGS and SoC for prospectively analyzed samples. **A)** Return time represents working days from receiving the tumor biopsy until a genomic report containing the diagnostically relevant molecular data was finalized. Primary results from SoC include combinations of various platform as deemed necessary in every case. Additional analyses (EPIC array or gene fusion panel) are labeled separately. **B)** Consumables cost estimation for every step of SoC molecular analysis of the prospectively analyzed samples. **C)** Schematic representation of how a combined approach incorporating nWGS and histological evaluation can be used to generate an integrated diagnostic report within five days. Panel C created in BioRender.

Hands-on time for a single nWGS sample was ∼three hours including DNA extraction from fresh/frozen tissue (30 minutes), library preparation (1.5 hours), flow cell loading (15 minutes) and washing/re-loading (45 minutes, optional). The hands-on time for SoC molecular analysis of the prospectively analyzed samples varied based on what type of analysis was deemed sufficient (minimum 11 hours, maximum 31 hours, Supplementary Table 7).

The reagent cost of analyzing a single sample by nWGS was estimated at 890 USD, which includes DNA extraction, library preparation and a single PromethION flow cell. The SoC-based median reagent cost for the prospectively analyzed samples was estimated at 1288 USD (minimum 273 USD, maximum 2157 USD, Figure 5B). This does not include the cost of labor and overhead.

## Discussion

We report that nWGS provides a comprehensive genetic work-up for CNS tumors in an uncomplicated, cost-effective pipeline. This approach enables not only the detection of all diagnostically relevant genetic alterations, comprising point mutations, SVs, gene fusions, and methylome-based tumor classification, but also potentially therapeutically relevant alterations, such as *MGMT* methylation status and therapeutic targets in a single assay (Figure 5C).

Precision oncology depends on molecular profiling for accurate tumor classification. A high fraction of biopsies is evaluated by complex molecular work-up - both for diagnostic clarity and identification of targeted therapy. Fougner et al describe a number of cases of GBM, IDHwt patients with actionable targets and therapeutic effects outside the standard treatment [21]. The 2021 WHO classification of CNS tumors requires DNA methylation profiling for specific entities [1, 22]. Similar requirements are increasingly observed across oncology, including carcinomas, sarcomas, and acute myeloid leukemia [4, 23, 24]. In this study, methylation-based classification from two of the three independent classifiers was concordant with SoC final diagnosis in 90% of cases, and in two cases nWGS provided methylation classifications that were subsequently confirmed by SoC and led to reclassification into newly defined CNS WHO entities. All diagnostically relevant alterations, including SNVs, CNVs, and gene fusions were consistently identified, in addition to complex homozygous *CDKN2A/B* deletions that were not found by FISH most likely due to probe size limitations. Our findings demonstrate that nWGS shows high concordance with SoC while providing additional diagnostic value through the detection of complex molecular variants and potential therapeutic targets not captured by common SoC NGS panels.

Turnaround time is a critical factor in neuro-oncology, where diagnosis-related treatment planning and trial enrolment depend on early molecular information. Conventional workflows frequently involve multiple assays, often performed sequentially over several weeks [25] delaying timely initiation of therapy and preclude trial enrolment. This prolongs hospitalization, increases workload for physicians, and escalates healthcare costs. Nanopore-based WGS addresses these limitations. This method captures both genetic and epigenetic alterations in a single workflow, avoiding fragmentation and tissue consumption inherent in conventional approaches. Without bisulfite conversion or RNA sequencing, it enables unbiased detection of clinically and diagnostically relevant alterations within a short timeframe. Importantly, the limited hands-on time and automated analytical pipeline reduce overall costs and enables a complete molecular work-up to be completed within few days.

Applying nanopore sequencing to precision cancer medicine has been undertaken by various groups over the past decade (reviewed in [26]). Methylation-based classification of CNS tumors has emerged as a particularly effective application, and multiple groups have demonstrated the feasibility of intraoperative CNS tumor diagnostics using nanopore sequencing [15, 27, 28]. Recently released workflows such as Rapid-CNS^2^ [16] and ROBIN [29] have combined intraoperative diagnostics with targeted sequencing of diagnostically relevant genomic regions. This approach produces rapid methylation-based classifications and detection of diagnostic variants at a cost of reduced sequencing output and computationally expensive and complex workflows. The reported median coverage of targeted regions after 24 hours of sequencing on PromethION flowcells was 30.4 [16] which is similar to nWGS coverage after 48 hours in this study (32.6x) but significantly lower than the median coverage of nWGS after 72 hours (44.5x). This may impact diagnostic performance, potentially resulting in insufficient sensitivity for fusions and SNVs. Furthermore, ONT’s rapid library protocol (∼10 minutes) is currently not applicable for targeted sequencing. This extends library preparation time which diminishes its utility in the intraoperative setting. For this reason, we have chosen to separate the intraoperative protocol from the nWGS full diagnostics protocol.

This study offers a practical comparison between nWGS results and those obtained through a typical SoC neuropathological workflow applied to real-world clinical samples. The SoC analysis aims to provide the minimal data necessary for diagnosis, prognostication, and treatment decisions. Consequently, many variants detected by nWGS lacked corresponding SoC data because they were not considered diagnostically significant for the given sample at the time. This is especially notable for SNVs, where nWGS findings were compared only to clinically relevant variants reported by SoC.

Nanopore-based WGS is not without limitations. In cases where the estimated TCC was below 15%, molecular findings, particularly SNV detection, proved unreliable. Variant calling of nanopore sequencing data has been shown to be comparable to short-read sequencing, with F1 accuracy reaching 99.99% [12]. However, the sensitivity limits of nWGS are higher than NGS panels due to lower sequencing depths. In this study, whole-genome sequencing depths ranged from 18.5–58x, compared to targeted panel sequencing that achieves much higher coverage in regions of interest. Although panel sequencing can detect low-frequency variants below the detection limit of nWGS, the clinical significance of such variants is uncertain, particularly in the absence of supporting histopathological or clonal evidence [30]. As with all molecular analysis, sufficient TCC is essential for representative nWGS results. Nanopore WGS is only possible from DNA extracted from fresh or frozen biopsies, which may be challenging in some cases. In this study, no case with TCC over 15% showed an inaccuracy that would have affected the final classification. To minimize the likelihood of false negative results due to sampling, the DIANA pipeline incorporates estimation of tumor cell content based on copy-number alterations. This allows cases with low TCC to be flagged and interpreted with caution, to assess whether findings are in line with the expected results or potentially require reinvestigation using complementary tools. Optimized tissue sampling and handling is imperative to secure diagnostic yield [31], as such all prospective cases contained adequate TCC.

Cost estimations and direct comparison between analytical platforms can be challenging. The cost of nWGS sequencing reagents in this study (DNA extraction, library prep and flow cells) was 890 USD per sample. A recent study [32] estimates the reagent cost of NGS panel sequencing for a single sample to be 1015 USD and the combined cost of consumables, labor, equipment financing and overhead to be 2944 USD per sample. Notably, this price per sample can only be obtained with optimal multiplexing (12 samples per run) and does not include methylation classification, structural variants or chromosomal aberrations. The cost of nWGS falls well within the average expenditure for NGS testing in an insurance financed healthcare system (Desai et al., 2021). In addition, reduction in turnaround time and hands-on work adds to the feasibility of nWGS in the diagnostical setting.

Molecular findings must be interpreted in conjunction with histological and immunohistochemical analyses, which typically require 3–5 workdays. Unlike targeted rapid assays, genome-wide nWGS can capture unexpected but diagnostically and clinically decisive events, ensuring diagnostic completeness. The complete genomic coverage obtained by nWGS enables the adaptation of previous diagnoses to updated WHO classification systems, as was observed in two cases of our retrospective cohort - one HGAP and one pHGG - where re-analysis by SoC confirmed the nWGS methylation classification findings and led to reclassification. While concordance between SoC and nWGS in this study was high, prospective validation in larger multi-center cohorts is warranted to confirm general clinical applicability.

In summary, nWGS consolidates comprehensive molecular diagnostics into a single assay that is accurate, scalable, and clinically feasible. By significantly shortening turnaround times while maintaining accuracy and expanding molecular insight, nWGS directly benefits patient care, supports timely therapeutic decisions, and offers a sustainable path forward for molecular neuro-oncology with potential for more efficient resource utilization at the healthcare system level.

## Conclusion

Integrating nWGS into routine diagnostics enables the simultaneous detection of diagnostically and therapeutically relevant genetic and epigenetic alterations. Our streamlined approach generates these results within a week, starting on the day of surgery, with limited hands-on time and costs comparable to current molecular work-up. By replacing multiple standalone analyses, including methylome profiling, MLPA, FISH and targeted NGS or fusion panels, nWGS offers a time- and cost-effective standardized solution for molecular diagnostics to improve both diagnostic accuracy and clinical decision-making.

## Data Availability

All data produced in the present study are available upon reasonable request to the authors

## Declaration of generative AI and AI-assisted technologies in the writing process

During the preparation of this work the authors used ChatGPT to correct grammar and increase readability. After using this tool, the authors reviewed and edited the content as needed and take full responsibility for the content of the publication.

## Supplementary figures

**Supplementary figure 1.**
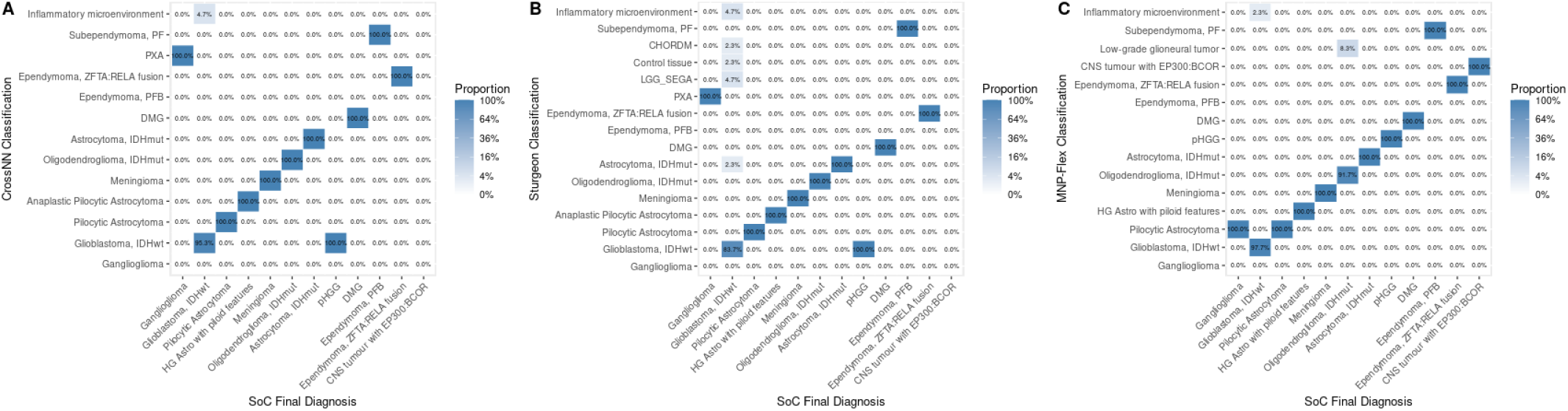
Methylation classifier accuracy confusion matrices of a) CrossNN, b) Sturgeon and c) MNP-Flex

**Supplementary figure 2.**
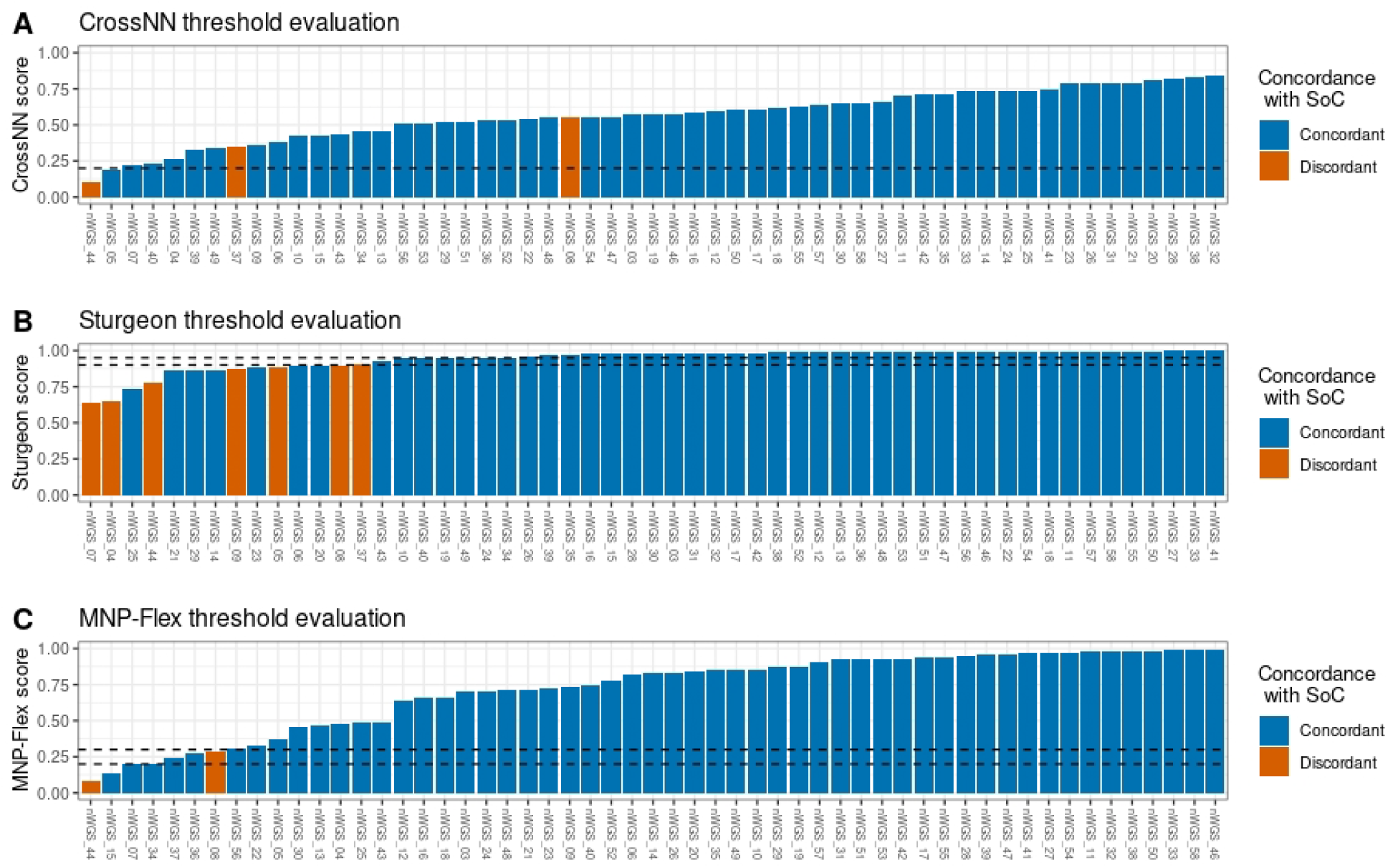
Threshold evaluation of methylation-based classifiers. Horizontal dotted lines represent suggested threshold values for each classifier. A) CrossNN, published high-confidence threshold = 0.2. B) Sturgeon, suggested thresholds are 0.95 (published, high-confidence) and 0.9. C) MNP-FLEX, published threshold for all platforms = 0.3, we suggest a threshold of 0.2 for nanopore whole-genome data. Discordant classification above threshold: nWGS_08 is GBM classified as “Inflammatory microenvironment” by all classifiers. nWGS_37 is pHGG classified GBM by CrossNN and Sturgeon.

**Supplementary Figure 3.**
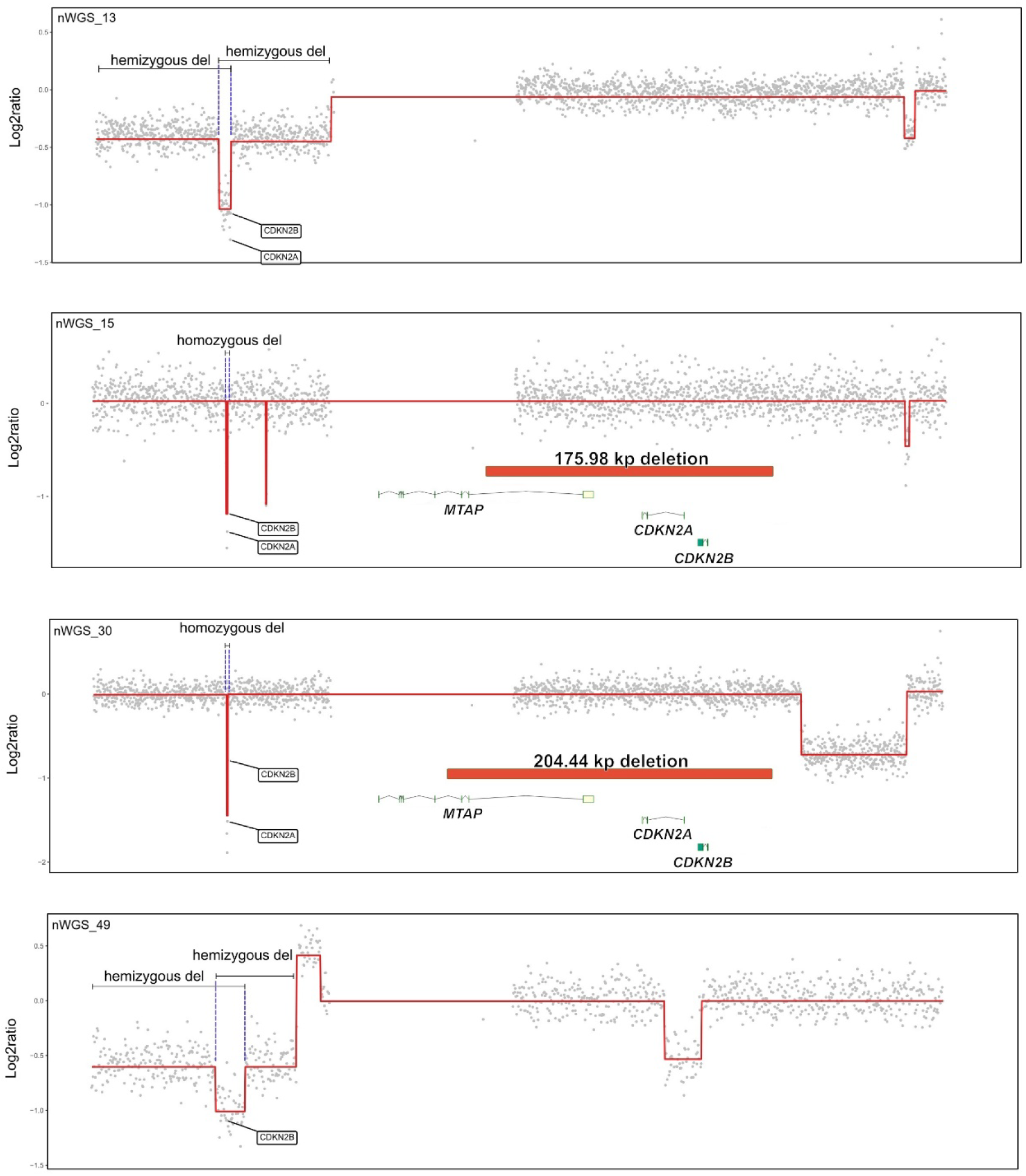
Discordant *CDKN2A/B* deletions between nWGS and SoC (FISH). nWGS CNV profiles indicated homozygous *CDKN2A/B* deletions in four samples (three GBM and an astrocytoma, IDH-mutant) where SoC (FISH) reported either no deletion (nWGS_13, nWGS_15 and nWGS_30) or a hemizygous deletion (nWGS_49). Overlapping hemizygous deletions were observed in two samples (nWGS_13 and nWGS_49), resulting in homozygous deletions at the *CDKN2A/B* loci. Homozygous deletions covering the *CDKN2A/B* loci were observed in the other two samples (nWGS_15 and nWGS_30). These homozygous deletions were confirmed by SV calling.

## References

1. Louis, D.N., et al., The 2021 WHO classification of tumors of the central nervous system: A summary. Neuro-Oncology, 2021. 23(8).

2. Capper, D., et al., EANO guideline on rational molecular testing of gliomas, glioneuronal, and neuronal tumors in adults for targeted therapy selection. Neuro-Oncology, 2023. 25(5): p. 813–826.

3. Silva, F.L.T., et al., Classification of pediatric soft and bone sarcomas using DNA methylation-based profiling. BMC cancer, 2024. 24(1).

4. Ren, M., et al., Mutational landscape and DNA methylation-based classification of squamous cell carcinoma and urothelial carcinoma. Clinical epigenetics, 2025. 17(1): p. 95–95.

5. Steinicke, T.L., et al., Rapid epigenomic classification of acute leukemia. Nature genetics, 2025.

6. Bertero, L., et al., Molecular neuropathology: an essential and evolving toolbox for the diagnosis and clinical management of central nervous system tumors, in Virchows Archiv. 2024.

7. Jagasia, S., et al., Cost Matrix of Molecular Pathology in Glioma-Towards AI-Driven Rational Molecular Testing and Precision Care for the Future. Biomedicines, 2022. 10(12).

8. Duncavage, E.J., et al., Genome Sequencing as an Alternative to Cytogenetic Analysis in Myeloid Cancers. New England Journal of Medicine, 2021. 384(10): p. 924–935.

9. Sosinsky, A., et al., Insights for precision oncology from the integration of genomic and clinical data of 13,880 tumors from the 100,000 Genomes Cancer Programme. Nature medicine, 2024. 30(1): p. 279–289.

10. Watkins, J.A., et al., Introduction and impact of routine whole genome sequencing in the diagnosis and management of sarcoma. British Journal of Cancer, 2024. 131(5): p. 860–869.

11. Meng, X., et al., Systematic evaluation of multiple NGS platforms for structural variants detection. Journal of Biological Chemistry, 2023. 299(12): p. 105436–105436.

12. Kolmogorov, M., et al., Scalable Nanopore sequencing of human genomes provides a comprehensive view of haplotype-resolved variation and methylation. Nature methods, 2023. 20(10): p. 1483–1492.

13. Bope, c.D., et al., DIANA: An integrated pipeline for analysis of long-read whole-genome sequencing data for molecular neuropathology. bioRxiv, 2026: p. 2026.03.25.714119.

14. Yuan, D., et al., crossNN is an explainable framework for cross-platform DNA methylation-based classification of tumors. Nature Cancer, 2025. 11: p. 1–12.

15. Vermeulen, C., et al., Ultra-fast deep-learned CNS tumour classification during surgery. Nature, 2023. 622(7984): p. 842–849.

16. Patel, A., et al., Prospective, multicenter validation of a platform for rapid molecular profiling of central nervous system tumors. Nature Medicine, 2025. 31(5): p. 1567–1577.

17. Broggi, G. and V. Barresi, Assessment of CDKN2A/B homozygous deletion in gliomas: to FISH or not to FISH? J Neuropathol Exp Neurol, 2023. 82(8): p. 742–744.

18. van den Bent, M.J., et al., Updated EANO guideline on rational molecular testing of gliomas, glioneuronal, and neuronal tumors in adults for targeted therapy selection - Update 1. Neuro-oncology, 2025. 2(27): p. 331–337.

19. Halldorsson, S., et al., Accurate and comprehensive evaluation of O6-methylguanine-DNA methyltransferase promoter methylation by nanopore sequencing. Neuropathology and applied neurobiology, 2024. 50(3).

20. Patel, A., et al., Rapid-CNS2: rapid comprehensive adaptive nanopore-sequencing of CNS tumors, a proof-of-concept study. Acta neuropathologica, 2022. 143(5): p. 609–612.

21. Fougner, V., et al., Actionable alterations in glioblastoma: Insights from the implementation of genomic profiling as the standard of care from 2016 to 2023. Neurooncol Pract, 2025. 12(1): p. 34–44.

22. Aldape, K., et al., cIMPACT-NOW update 9: Recommendations on utilization of genome-wide DNA methylation profiling for central nervous system tumor diagnostics. Neuro-oncology advances, 2025. 7(1).

23. Khan, A.A., et al., Sarcoma diagnosis by DNA methylation classifier: A systematic review, current status and future prospects. Pathology, research and practice, 2024. 263: p. 155634–155634.

24. Marchi, F., et al., Rapid Long-Read Epigenomic Diagnosis and Prognosis of Acute Myeloid Leukemia. Blood, 2024. 144(Supplement 1): p. 103–103.

25. Navani, N., et al., Optimising tissue acquisition and the molecular testing pathway for patients with non-small cell lung cancer: A UK expert consensus statement. Lung cancer (Amsterdam, Netherlands), 2022. 172: p. 142–153.

26. Dyshlovoy, S.A., et al., Applications of Nanopore sequencing in precision cancer medicine. International Journal of Cancer, 2024. 155(12): p. 2129–2140.

27. Brändl, B., et al., Rapid brain tumor classification from sparse epigenomic data. Nature Medicine, 2025: p. 1–9.

28. Djirackor, L., et al., Intraoperative DNA methylation classification of brain tumors impacts neurosurgical strategy. Neuro-Oncology Advances, 2021. 3(1).

29. Deacon, S., et al., ROBIN: A unified nanopore-based assay integrating intraoperative methylome classification and next-day comprehensive profiling for ultra-rapid tumor diagnosis. Neuro-Oncology, 2025.

30. Li, M.M., et al., Standards and Guidelines for the Interpretation and Reporting of Sequence Variants in Cancer: A Joint Consensus Recommendation of the Association for Molecular Pathology, American Society of Clinical Oncology, and College of American Pathologists. Journal of Molecular Diagnostics, 2017. 19(1): p. 4–23.

31. Karschnia, P., et al., A framework for standardised tissue sampling and processing during resection of diffuse intracranial glioma: joint recommendations from four RANO groups. Lancet Oncol, 2023. 24(11): p. e438–e450.

32. Henkel, P.S., et al., Microcosting Study of Genomic Profiling for Precision Cancer Medicine: Application from the National Infrastructure for Precision Diagnostics in Norway. J Mol Diagn, 2025. 27(10): p. 945–953.

